# Multi-Ancestry Survival GWAS of Substance Use Initiation in the ABCD Study

**DOI:** 10.64898/2026.04.08.26350431

**Authors:** Mengman Wei, Qian Peng

**Affiliations:** Department of Neuroscience, The Scripps Research Institute, 10550 N Torrey Pines Rd, La Jolla, 92037, CA, U.S.

**Keywords:** survival GWAS, meta-analysis, ABCD, substance use initiation, nicotine, cannabis, alcohol, multi-ancestry

## Abstract

**Background:** Substance use initiation in adolescence is influenced by both genetic and environmental factors; however, large-scale genetic studies often treat initiation as a binary outcome and underuse longitudinal timing information.

**Methods:** We conducted time-to-event (survival) genome-wide association analyses (GWAS) of initiation for four outcomes—alcohol, nicotine, cannabis, and any substance use—using longitudinal follow-up data from the Adolescent Brain Cognitive Development (ABCD) Study. We performed ancestry-stratified GWAS within European (EUR), African (AFR), and Hispanic (HISP) groups, applying consistent quality control and covariate adjustment. Summary statistics were harmonized across ancestries and meta-analyzed using inverse-variance weighted fixed-effects and DerSimonian–Laird random-effects models. We evaluated genomic inflation and heterogeneity (Cochran’s *Q* and *I*^2^), identified independent lead variants at genome-wide and suggestive significance thresholds, and assessed cross-trait overlap of associated loci.

**Results:** In the multi-ancestry meta-analysis, we observed suggestive association signals across traits (minimum *p*-values: alcohol ∼ 1 × 10^−7^, any ∼ 1 × 10^−7^, cannabis ∼ 5 × 10^−8^, nicotine ∼ 1 × 10^−8^). Nicotine initiation showed one genome-wide significant variant in both fixed- and random-effects meta-analyses (*p* < 5 × 10^−8^). Across traits, suggestive loci demonstrated limited overlap, with the strongest concordance between alcohol and any substance use, consistent with shared liability. Heterogeneity statistics indicated that some loci exhibited cross-ancestry variation in effect estimates.

**Conclusions:** Survival GWAS leveraging initiation timing can identify genetic signals that may be missed by binary designs and enables principled multi-ancestry synthesis. Our results highlight both shared and trait-specific genetic contributions to early substance initiation and provide a foundation for downstream functional annotation and integrative modeling with environmental risk factors. These findings demonstrate the value of incorporating developmental timing into genetic discovery and provide a framework for integrating longitudinal risk modeling with genomic analyses.

## Introduction

Early initiation of substance use is associated with elevated risk of later substance use disorders and adverse psychiatric outcomes [1, 2, 3]. Genetic factors contribute substantially to liability for substance use behaviors, yet discovery in youth cohorts remains challenging because initiation is strongly age-dependent, follow-up is incomplete, and event rates vary across substances [4, 5, 6]. Many GWAS of initiation also collapse longitudinal information into a binary phenotype (e.g., ever/never initiation), which can reduce power and obscure timing-related effects [6, 7].

Time-to-event modeling offers a principled alternative by incorporating both events (initiation) and censoring (no initiation by last follow-up), thus using more information from longitudinal cohorts [8, 9, 10]. The Adolescent Brain Cognitive Development (ABCD) Study provides repeated assessments across adolescence, enabling survival GWAS of initiation across multiple substances and an “any initiation” composite outcome [11, 12, 13]. We hypothesized that incorporating time-to-initiation information would increase statistical power relative to binary GWAS and reveal both shared and substance-specific genetic architectures across ancestries.

Here, we report a multi-ancestry survival GWAS of initiation for alcohol, nicotine, cannabis, and any substance use in ABCD. We conduct ancestry-stratified GWAS in European (EUR), African (AFR), and Hispanic/Latino (HISP) groups and perform a strand-aware meta-analysis across ancestries using fixed- and random-effects models [14, 15, 16, 17]. Finally, we compare cross-trait overlap of suggestive loci to quantify shared versus substance-specific genetic architecture [6].

To our knowledge, this is among the first studies to apply survival GWAS to substance use initiation in a longitudinal adolescent cohort and to integrate results across multiple ancestries using a strand-aware meta-analysis framework.

## Methods

### Study cohort

The Adolescent Brain Cognitive Development (ABCD) Study [13] is a longitudinal study that examines behavioral and biological development from middle childhood to young adulthood. At baseline, 11,875 children aged 8, 9 to 11 years (born 2005–2009) were recruited from 22 research sites across the United States. The study protocol was approved by the institutional review board at each site. Race and ethnicity were reported by parents or caregivers to describe the sociodemographic characteristics of the sample. All data used in this study were obtained from the ABCD Study release 5.1. We used ABCD participants with genotype data passing quality control and at least one valid follow-up assessment for substance use initiation. Participants were assigned to ancestry groups (EUR/AFR/HISP) using ABCD-provided ancestry labels, and analyses were restricted to individuals within each ancestry group where applicable.

### Phenotypes: time-to-initiation outcomes

For each outcome (alcohol, nicotine, cannabis, and any substance), we created time-to-event (survival) measures of initiation. Time was defined as the number of months from the baseline visit to either the first report of substance initiation (event) or the last available follow-up visit (censored). The event was coded as 1 if the participant reported first use during follow-up and 0 if they had no reported use by their last observed visit. So time-to-event was defined as months since baseline assessment, with baseline age included as a covariate.

We applied the same initiation criteria across all study visits, and we defined censoring time as the final assessment at which valid substance use reporting was available for the participant. This approach matches the substance use initiation definitions and censoring rules used in a recent ABCD substance initiation analysis study [18].

### Genotype Data and Quality Control

Genotype data from the ABCD Study were processed using PLINK v1.9 [19, 20]. We used the imputed genotype dataset provided by ABCD and applied standard quality control procedures to both samples and variants.

First, we restricted the genotype dataset to individuals included in the curated ABCD genotype sample list. Sex information from the curated genotype dataset was used to update the imputed genotype files to ensure consistency between genotype and phenotype records.

We then applied sample-level quality control. Individuals with genotype missingness greater than 5% were removed (`--mind 0.05`). Next, we applied variant-level quality control filters. Variants were excluded if they met any of the following criteria: minor allele frequency (MAF) < 0.01; genotype missingness > 5% (`--geno 0.05`); Hardy–Weinberg equilibrium *p*-value < 1 × 10^−6^ (`--hwe 1e-6`).

After these filters, the remaining variants were retained for downstream analyses. For analyses requiring linkage disequilibrium (LD) independence, we additionally performed LD pruning using a sliding window approach (`--indep-pairwise 50 5 0.2`). The pruned set was used for LD-based analyses such as principal component analysis, while the full QC-filtered dataset was used for genome-wide association analyses.

### Additional Variant Filtering for Survival GWAS

Before performing survival GWAS, additional variant filters were applied within each ancestry group to ensure stable model estimation.

Variants were restricted to autosomal chromosomes (1–22). We applied the following filters: variant missingness ≤ 2% (`--geno 0.02`); minor allele count (MAC) ≥ 10 for Step-1 model training; Hardy– Weinberg equilibrium *p* ≥ 1×10^−6^ within each ancestry group; SNPs restricted to standard A/C/G/T alleles; and exclusion of variants located in known long-range linkage disequilibrium (LD) regions (e.g., the major histocompatibility complex (MHC) region on chromosome 6).

Variants passing these filters were further LD-pruned(`--indep-pair wise 1000 100 0.2`) to construct the SNP set used for the REGENIE [21] Step-1 prediction model. In Step-2 association testing, variants were required to have minor allele count ≥ 20 to ensure stable effect estimation. Step-1 SNP selection and LD pruning were performed using unrelated individuals, whereas model fitting and association testing were conducted using all individuals within each ancestry group.

### Covariates

All association models included the following covariates: gender; baseline age; study site (categorical); the first 20 genetic principal components (PCs) to account for residual population structure. Principal components were calculated from genotype data within each ancestry group.

### Ancestry-stratified survival GWAS

Cox proportional hazards models were fitted using REGENIE 4.1 [21], which implements a two-step whole-genome regression framework optimized for large datasets, with Efron approximation for tied event times. The proportional hazards assumption was evaluated using Schoenfeld residuals in a subset of top loci.

#### Step 1: Polygenic model fitting

In Step-1, a ridge regression model was fitted using a set of LD-pruned variants to estimate polygenic predictions while accounting for relatedness and polygenic background effects. Five-fold cross-validation was used to generate leave-one-chromosome-out (LOCO) predictions.

#### Step 2: Association testing

In Step-2, genome-wide association tests were performed using the LOCO predictions from Step-1 as offsets. Each variant was tested using a Cox proportional hazards model [8, 22] implemented in REGENIE. Variants with minor allele count < 20 were excluded in Step-2 association testing. Analyses were conducted separately for each ancestry group and for each substance initiation phenotype.

### Multi-ancestry meta-analysis

Meta-analysis was performed to combine ancestry-specific survival GWAS results across European (EUR), African (AFR), and Hispanic (HISP) ancestry groups. Prior to meta-analysis, variants were harmonized across datasets using the European ancestry results as the reference. Alleles were aligned across studies, and effect sizes were sign-flipped when necessary to ensure consistent orientation. Palindromic variants with allele frequencies close to 0.5 were excluded to avoid strand ambiguity.

Association statistics were combined using inverse-variance weighted fixed-effects meta-analysis. Heterogeneity across ancestry groups was assessed using Cochran’s Q statistic [23] and the I^2^ metric [24]. Random-effects estimates were additionally calculated using the DerSimonian–Laird method [25]. Meta-analysis was conducted separately for each substance initiation phenotype (alcohol, nicotine, cannabis, and any substance). All meta-analyses were implemented using R version 4.3.0 meta / metafor package [26].

### Functional annotation

Functional annotation of meta-analysis results was performed using FUMA (v1.8.2) [27, 28]. Independent significant SNPs (*p* < 1×10^−6^) and lead SNPs (*r*^2^ < 0.1) were defined using the 1000 Genomes Project Phase 3 reference panel [29]. SNPs were mapped to genes using positional mapping (*±*10 kb), expression quantitative trait loci (eQTL) mapping based on GTEx v8 across all tissues [30], and chromatin interaction mapping. Gene-based analysis and tissue enrichment were performed using MAGMA [31] as implemented in FUMA.

## Results

### GWAS quality control

The initial imputed genotype dataset contained 11,666 individuals and 8,118,068 variants after genotype quality control. Individuals were stratified by genetic ancestry and analyzed separately for European (EUR), African (AFR), and Hispanic (HISP) ancestry groups.

In the European ancestry group, 5,476 individuals were available for analysis, including 2,388 alcohol initiation events, 2,507 events for any substance initiation, 183 cannabis initiation events, and 286 nicotine initiation events. A subset of 4,683 unrelated individuals was identified and used for the REGENIE Step-1 model fitting.

In the African ancestry group, 1,323 individuals were included in the analysis, with 282 alcohol initiation events, 362 events for any substance initiation, 60 cannabis initiation events, and 64 nicotine initiation events. Among these, 996 unrelated individuals were used for model training.

In the Hispanic ancestry group, 1,995 individuals were included, with 685 alcohol initiation events, 773 events for any substance initiation, 86 cannabis initiation events, and 134 nicotine initiation events. A total of 1,343 unrelated individuals were used in the Step-1 model fitting.

The unrelated subsets were used to construct the ridge regression model in REGENIE Step-1, while association testing in Step-2 was conducted using all available individuals within each ancestry group.

After variant-level quality control, 6,530,186 variants in the European ancestry dataset passed filtering criteria. Linkage disequilibrium (LD) pruning (*r*^2^ < 0.2) resulted in 416,861 approximately independent variants, which were used to fit the REGENIE Step-1 ridge regression model.

In the African ancestry dataset, 7,377,739 variants passed quality control, of which 43,992 variants remained after LD pruning. In the Hispanic ancestry dataset, 7,572,579 variants passed variant filtering, and 43,980 variants were retained after LD pruning.

These LD-pruned variants were used to estimate polygenic predictions in REGENIE Step-1, which were subsequently used as offsets in the Step-2 genome-wide association analyses.

### Genome-wide association analyses of substance initiation

We conducted survival GWAS for alcohol, nicotine, cannabis, and any substance initiation in the ABCD cohort, followed by multi-ancestry meta-analysis across EUR, AFR, and HISP populations. Gene-based and gene-set analyses were corrected for multiple testing using Bonferroni/FDR correction as implemented in MAGMA. Details of the results could be seen in the supplementary.

#### GWAS calibration and polygenicity

Genome-wide test statistics were well calibrated across all traits, with modest inflation observed in the full sample combining European (EUR), African (AFR), and Hispanic (HISP) populations (λ_GC_ ≈ 1.08–1.13; mean *χ*^2^ ≈ 1.08–1.14), consistent with polygenic traits in modest sample sizes. LD score regression intercepts were close to 1, indicating that the observed inflation was primarily attributable to polygenicity rather than confounding.

Across traits, alcohol and any substance initiation showed slightly stronger signal (λ_GC_ ≈ 1.13), whereas nicotine and cannabis showed more modest inflation (λ_GC_ ≈ 1.08–1.11), consistent with differences in event rates and statistical power. Effect sizes of lead variants were modest (hazard ratios approximately 1.05–1.15 per allele), consistent with a highly polygenic architecture typical of behavioral traits.

#### Cross-ancestry patterns

Ancestry-specific analyses revealed differences in signal strength across populations. For all traits, the strongest signal was observed in the European (EUR) sample (λ_GC_ ≈ 1.04–1.13), reflecting its larger effective sample size.

For alcohol and any substance initiation, association signals were relatively consistent across ancestries, with moderate inflation observed in African (AFR) and Hispanic (HISP) populations (λ_GC_ ≈ 1.05–1.06).

In contrast, nicotine and cannabis showed reduced signal in AFR (λ_GC_ < 1) and moderate inflation in HISP (λ_GC_ ≈ 1.06–1.09), suggesting that meta-analysis results for these traits are primarily driven by the EUR sample, likely due to differences in sample size and linkage disequilibrium (LD) structure across populations.

#### Meta-analysis and downstream functional annotation

Given the increased statistical power of the combined analysis, downstream functional annotation was performed using the meta-analysis results. Gene mapping and gene-set enrichment analyses prioritized candidate genes and pathways, highlighting transcriptional regulation, immune-related signaling, and developmental gene clusters (e.g., HOXA genes) as potential biological mechanisms underlying substance initiation.

### Multi-ancestry meta-analysis discovery

Fixed-effects meta-analysis combining EUR, AFR, and HISP ancestry-stratified survival GWAS results identified several suggestive associations across substance initiation phenotypes. The smallest fixed-effects meta-analysis *p*-values were 1.07 × 10^−7^ for alcohol initiation, 1.19 × 10^−7^ for any substance initiation, 5.53 × 10^−8^ for cannabis initiation, and 1.38 × 10^−8^ for nicotine initiation.

At the suggestive threshold of *p* < 1 × 10^−6^, there were 30 associated variants for alcohol initiation, 15 for any substance initiation, 21 for cannabis initiation, and 11 for nicotine initiation. At *p* < 1 × 10^−5^, the corresponding numbers were 208, 196, 151, and 150, respectively.

Only the nicotine initiation meta-analysis yielded a genome-wide significant association under the conventional threshold of *p* < 5 × 10^−8^. Cannabis initiation showed a near-threshold association (minimum *p* = 5.53 × 10^−8^), whereas alcohol initiation and any substance initiation showed multiple suggestive but not genome-wide significant signals, as shown in Table 2.

**Table 1.**
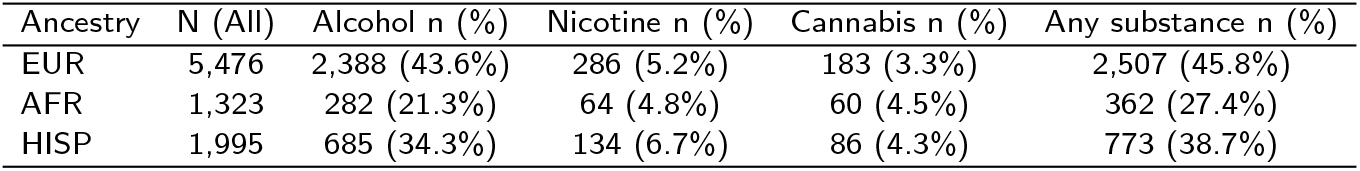
Sample sizes by ancestry group and substance initiation outcomes.

| Ancestry | N (All) | Alcohol n (%) | Nicotine n (%) | Cannabis n (%) | Any substance n (%) |
| --- | --- | --- | --- | --- | --- |
| EUR | 5,476 | 2,388 (43.6%) | 286 (5.2%) | 183 (3.3%) | 2,507 (45.8%) |
| AFR | 1,323 | 282 (21.3%) | 64 (4.8%) | 60 (4.5%) | 362 (27.4%) |
| HISP | 1,995 | 685 (34.3%) | 134 (6.7%) | 86 (4.3%) | 773 (38.7%) |

**Table 2.** Top independent lead loci from the multi-ancestry survival GWAS meta-analysis of alcohol, nicotine, cannabis, and any substance initiation in the ABCD Study.

| Trait | CHR | BP | SNP | A1 | A2 | Beta | SE | HR | CI<sub>lower</sub> | CI<sub>upper</sub> | $p$ | N | $k$ | $I^2$ |
| --- | --- | --- | --- | --- | --- | --- | --- | --- | --- | --- | --- | --- | --- | --- |
| alcohol | 13 | 42608671 | rs9567001 | G | A | -0.6079 | 0.1144 | 0.544 | 0.435 | 0.681 | $1.073 \times 10^{-7}$ | 8,794 | 3 | 0 |
| alcohol | 2 | 118329078 | chr2:118329078 | C | T | -0.2366 | 0.0447 | 0.789 | 0.723 | 0.862 | $1.226 \times 10^{-7}$ | 8,794 | 3 | 27.74 |
| alcohol | 2 | 45972216 | rs2166969 | T | C | 0.1565 | 0.0298 | 1.169 | 1.103 | 1.240 | $1.48 \times 10^{-7}$ | 8,794 | 3 | 70.23 |
| alcohol | 8 | 5247565 | rs74428965 | T | C | 0.2423 | 0.0475 | 1.274 | 1.161 | 1.398 | $3.392 \times 10^{-7}$ | 8,794 | 3 | 0 |
| alcohol | 6 | 1309248 | rs74940956 | T | G | -0.2663 | 0.0528 | 0.766 | 0.691 | 0.850 | $4.516 \times 10^{-7}$ | 8,794 | 3 | 0 |
| nicotine | 9 | 24973434 | chr9:24973434 | A | AT | 1.7815 | 0.3139 | 5.939 | 3.210 | 10.987 | $1.379 \times 10^{-8}$ | 7,471 | 2 | 0 |
| nicotine | 9 | 137699491 | rs144112581 | A | G | 1.4684 | 0.2781 | 4.342 | 2.517 | 7.490 | $1.298 \times 10^{-7}$ | 7,471 | 2 | 0 |
| nicotine | 5 | 85736434 | chr5:85736434 | C | CTTAA | 1.4096 | 0.2721 | 4.094 | 2.402 | 6.980 | $2.224 \times 10^{-7}$ | 7,471 | 2 | 54.11 |
| nicotine | 5 | 127713402 | rs78607920 | C | A | 1.5936 | 0.3080 | 4.921 | 2.691 | 9.001 | $2.294 \times 10^{-7}$ | 8,794 | 3 | 91.50 |
| nicotine | 7 | 139059933 | rs74348184 | C | T | 1.1889 | 0.2323 | 3.283 | 2.082 | 5.177 | $3.093 \times 10^{-7}$ | 8,794 | 3 | 13.24 |
| cannabis | 6 | 148006492 | rs17077870 | G | C | 0.5694 | 0.1048 | 1.767 | 1.439 | 2.170 | $5.534 \times 10^{-8}$ | 7,471 | 2 | 0 |
| cannabis | 2 | 18437017 | rs16985014 | T | C | 1.7247 | 0.3195 | 5.611 | 2.999 | 10.495 | $6.742 \times 10^{-8}$ | 7,471 | 2 | 87.10 |
| cannabis | 16 | 86123749 | rs147860977 | C | T | 1.5701 | 0.2957 | 4.807 | 2.693 | 8.581 | $1.094 \times 10^{-7}$ | 7,471 | 2 | 0 |
| cannabis | 16 | 8625400 | rs75552258 | A | G | 1.9317 | 0.3690 | 6.902 | 3.349 | 14.224 | $1.646 \times 10^{-7}$ | 7,471 | 2 | 10.61 |
| cannabis | 1 | 53078841 | rs12567021 | T | C | 1.4595 | 0.2814 | 4.304 | 2.479 | 7.471 | $2.139 \times 10^{-7}$ | 8,794 | 3 | 0 |
| any | 6 | 1309248 | rs74940956 | T | G | -0.2708 | 0.0511 | 0.763 | 0.690 | 0.843 | $1.186 \times 10^{-7}$ | 8,794 | 3 | 0 |
| any | 16 | 14589571 | chr16:14589571 | G | GA | -0.5120 | 0.0972 | 0.599 | 0.495 | 0.725 | $1.4 \times 10^{-7}$ | 7,471 | 2 | 0 |
| any | 11 | 117436522 | chr11:117436522 | A | G | -0.3028 | 0.0591 | 0.739 | 0.658 | 0.830 | $3.043 \times 10^{-7}$ | 8,794 | 3 | 78.17 |
| any | 2 | 45972216 | rs2166969 | T | C | 0.1456 | 0.0287 | 1.157 | 1.093 | 1.224 | $4.108 \times 10^{-7}$ | 8,794 | 3 | 53.20 |
| any | 8 | 5067910 | rs13265017 | A | T | -0.1473 | 0.0291 | 0.863 | 0.815 | 0.914 | $4.193 \times 10^{-7}$ | 7,471 | 2 | 4.65 |

Using a locus definition based on lead variants with *p* < 1 × 10^−5^ and a *±*250 kb window, we identified 52 lead loci for alcohol initiation, 51 lead loci for any substance initiation, 61 lead loci for cannabis initiation, and 53 lead loci for nicotine initiation. Full results are provided in the Supplementary Materials.

### Heterogeneity across ancestries

Between-ancestry heterogeneity was assessed using Cochran’s *Q* statistic and the *I*^2^ metric. Across all four traits, the median *I*^2^ was 0, indicating that most variants showed little or no detectable heterogeneity across ancestry groups.

For alcohol initiation, 413,290 of 6,750,735 variants (6.12%) showed nominal heterogeneity (*Q*-test *p* < 0.05), 1,070,437 (15.86%) had *I*^2^ > 50%, and 196,314 (2.91%) had *I*^2^ > 75%. For any substance initiation, the corresponding values were 412,313 (6.11%), 1,070,958 (15.86%), and 193,602 (2.87%).

For nicotine initiation, 330,998 variants (4.90%) had *Q*-test *p* < 0.05, 907,747 (13.45%) had *I*^2^ > 50%, and 157,829 (2.34%) had *I*^2^ > 75%. For cannabis initiation, 315,703 variants (4.68%) had *Q*-test *p* < 0.05, 885,291 (13.11%) had *I*^2^ > 50%, and 146,537 (2.17%) had *I*^2^ > 75%.

Overall, these results indicate that the majority of meta-analyzed variants had consistent effects across ancestries, although a subset of loci showed moderate-to-high heterogeneity.

### Cross-trait overlap and concordance

Comparison of lead loci across traits revealed the greatest overlap between alcohol initiation and any substance initiation. Of the 52 alcohol lead loci and 51 any-substance lead loci, 29 loci were shared, corresponding to a Jaccard index of 0.392. Among these shared loci, 96.6% showed concordant directions of effect.

In contrast, there was no lead-locus overlap between cannabis initiation and either alcohol initiation or any substance initiation. Overlap between nicotine initiation and the other phenotypes was limited: 2 shared loci with alcohol initiation (Jaccard index = 0.019) and 3 shared loci with any substance initiation (Jaccard index = 0.030), with direction concordance proportions of 0.50 and 0.33, respectively.

Cannabis initiation and nicotine initiation shared 2 loci (Jaccard index = 0.018), both of which showed concordant effect directions.

Taken together, these findings suggest a substantial shared genetic signal between alcohol initiation and broader substance initiation, but comparatively limited overlap involving cannabis or nicotine initiation, consistent with partially shared liability pathways rather than coincidental overlap.

### Functional Annotation and Biological Interpretation

For nicotine initiation, the multi-ancestry meta-analysis of survival GWAS results identified one genome-wide significant locus. The strongest association was observed on chromosome 9 at SNP rs42974334, with a meta-analysis effect estimate of *β* = 1.022, corresponding to a hazard-ratio estimate of approximately exp(1.022) ≈ 2.78, and a genome-wide significant association signal (*P* = 1.38 × 10^−8^). This result suggests that genetic variation at or near this locus may be associated with increased hazard of nicotine initiation during adolescence.

To evaluate whether this association pattern was specific to time-to-event modeling, we compared the full-sample binary GWAS and survival GWAS results for nicotine initiation. The primary binary GWAS did not identify any genome-wide significant loci, with the strongest association observed at rs114790912 on chromosome 8 (*P* = 1.48 × 10^−6^). In contrast, the full-sample survival GWAS identified a stronger genome-wide significant signal at rs42974334 on chromosome 9 (*P* = 1.31×10^−10^). This contrast suggests that modeling the timing of nicotine initiation may capture additional genetic signal not fully represented by a cross-sectional binary phenotype.

**Figure 1.**
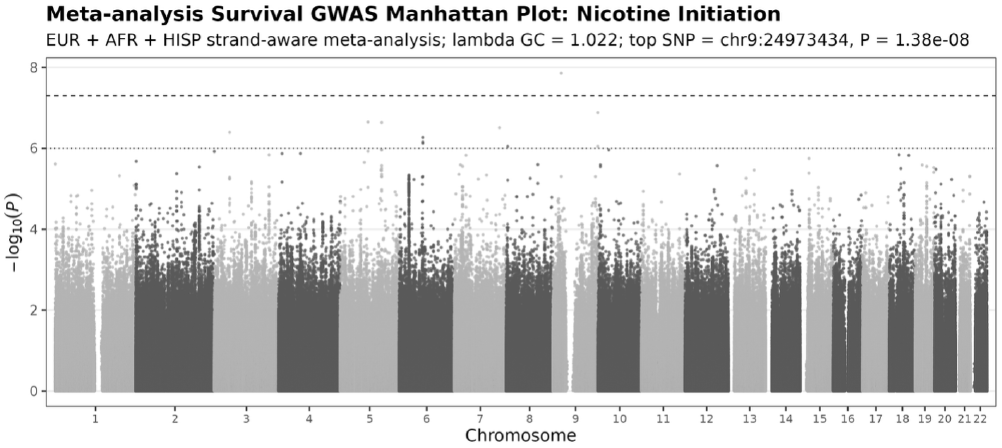
Multi-ancestry meta-analysis survival GWAS Manhattan plot for nicotine initiation. The plot shows chromosome-wide association results from the EUR, AFR, and HISP ancestry-stratified survival GWAS meta-analysis. Each point represents one SNP, with genomic position shown on the x-axis and − log_10_(*P*) shown on the y-axis. The dashed horizontal line indicates the conventional genome-wide significance threshold of *P* < 5 × 10^−8^. The top signal was observed at rs42974334 on chromosome 9, with a meta-analysis effect estimate of *β* = 1.022 and *P* = 1.38 × 10^−8^.

The stronger association observed in the survival GWAS may reflect the additional information provided by age or follow-up timing of first nicotine use. Rather than treating all initiators equivalently, the time-to-event model accounts for when initiation occurred and incorporates censoring information from participants who had not initiated nicotine use by their last available follow-up. This framework may therefore provide greater sensitivity for detecting genetic effects associated with earlier nicotine initiation.

To support biological interpretation of the chromosome 9 locus, we next performed functional annotation using positional mapping, gene-based annotation, and tissue-enrichment analyses. The lead SNP and nearby variants were mapped to candidate genes based on genomic proximity and, where available, regulatory annotations. We further evaluated whether genes near the associated locus were enriched in biologically relevant tissues, including brain regions and tissues involved in neurodevelopment, reward processing, and addiction vulnerability.

**Figure 2.**
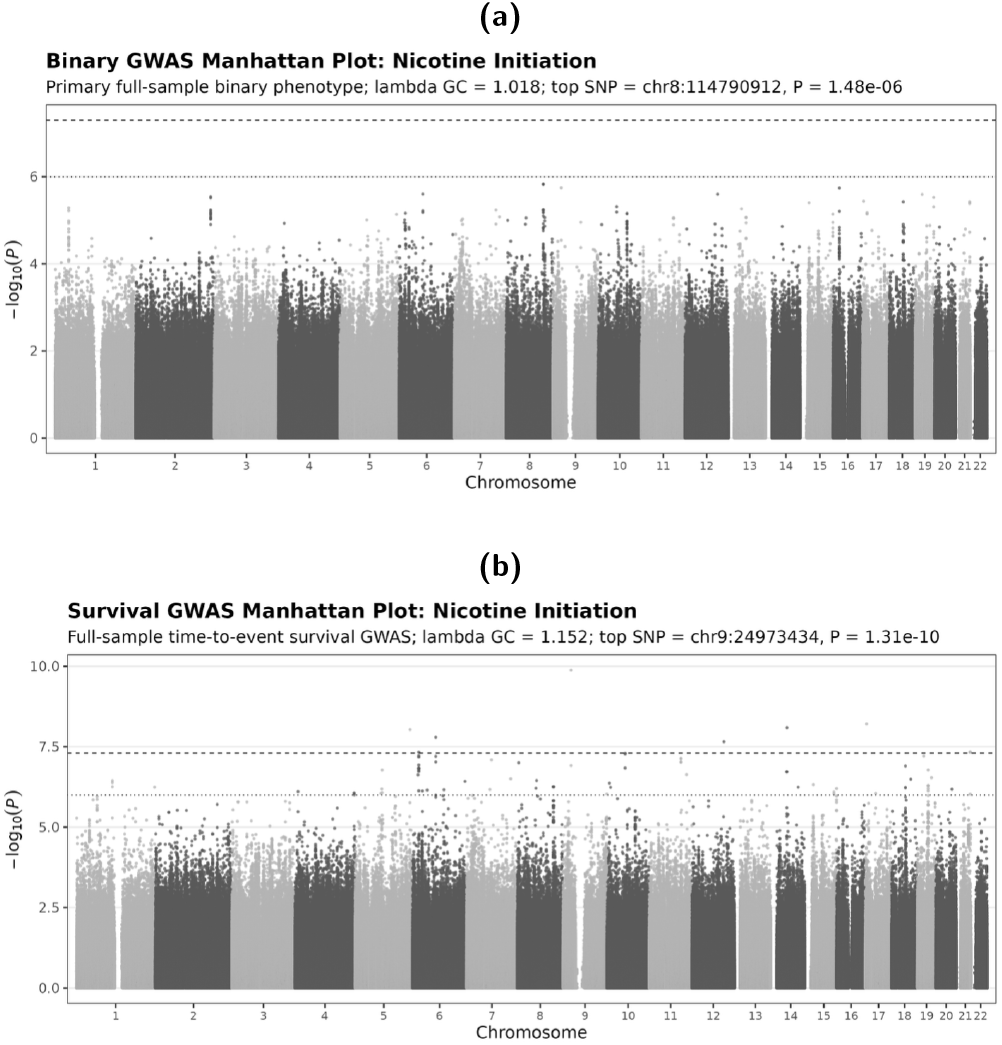
Comparison of binary and survival GWAS results for nicotine initiation. **(a)** Manhattan plot from the primary full-sample binary GWAS of nicotine initiation. The strongest association was observed at rs114790912 on chromosome 8, with genomic-control inflation factor λ_GC_ = 1.018 and *P* = 1.48 × 10^−6^, which did not reach the conventional genome-wide significance threshold. **(b)** Manhattan plot from the full-sample time-to-event survival GWAS of nicotine initiation. The survival GWAS showed a genome-wide significant signal at rs42974334 on chromosome 9, with λ_GC_ = 1.152 and *P* = 1.31×10^−10^. The dashed horizontal line indicates the conventional genome-wide significance threshold of *P* < 5 × 10^−8^.

Although the chromosome 9 signal reached genome-wide significance, interpretation should remain cautious. Nicotine initiation events are relatively sparse in the ABCD cohort compared with continuous behavioral outcomes, and the survival GWAS showed higher genomic inflation (λ_GC_ = 1.152) than the binary GWAS (λ_GC_ = 1.018). Therefore, this locus should be treated as a promising candidate signal rather than a definitive mechanistic finding. Replication in larger adolescent or longitudinal substance-use cohorts will be important to confirm the robustness of this association and to determine whether the locus contributes specifically to nicotine initiation or more broadly to externalizing and substance-use liability.

Overall, the nicotine initiation GWAS suggests a potentially meaningful genetic contribution to early nicotine-use risk, while also highlighting the need for careful downstream annotation and replication. These results provide a basis for further investigation of chromosome 9 candidate genes and their possible roles in adolescent risk-taking, reward sensitivity, and vulnerability to substance initiation.

## Discussion

In this study, we leveraged longitudinal data from the ABCD cohort to perform survival genome-wide association studies (GWAS) of substance use initiation, followed by a strand-aware multi-ancestry meta-analysis across European (EUR), African (AFR), and Hispanic (HISP) ancestry groups. By incorporating time-to-event information and censoring, the survival GWAS framework makes more efficient use of the available data compared to traditional binary approaches. In addition, the strand-aware harmonization strategy reduces allele alignment errors, enabling more reliable integration of results across ancestries.

Although we observed limited genome-wide significant associations for alcohol, cannabis, and overall substance initiation, we identified consistent suggestive signals across traits, as well as a genome-wide significant locus for nicotine initiation. This pattern is expected in adolescent cohorts such as ABCD, where relatively low event rates and limited follow-up duration constrain statistical power. Furthermore, although multi-ancestry meta-analysis improves generalizability, it may attenuate association signals when genetic effects differ across populations or when linkage disequilibrium (LD) patterns vary. Compared with large-scale adult GWAS [6], the effect sizes observed here are smaller, which is consistent with the developmental stage of the cohort. Genetic influences on substance use may become more pronounced later in life, whereas early adolescence is likely more strongly shaped by environmental factors. Nevertheless, these findings highlight the potential for early risk prediction and inform prevention strategies targeting modifiable factors during critical developmental windows.

In addition, this study focuses on substance initiation during early adolescence, a developmental period characterized by rapid neurobiological and behavioral changes. The relatively modest genetic signals observed here are consistent with the notion that environmental and developmental factors may exert stronger influence at this stage, with genetic effects becoming more pronounced later in life.

Importantly, our results highlight the advantages of survival GWAS over traditional case–control GWAS. By modeling the timing of initiation events, survival GWAS captures additional information about disease development that is not available in binary phenotypes. Consistent with this, parallel analyses using conventional binary GWAS on the same phenotypes did not yield significant findings in the ABCD dataset. These results suggest that survival-based approaches can improve power, particularly in moderately sized cohorts such as ABCD, where incorporating temporal information can help uncover genetic signals that would otherwise remain undetected.

This study should be viewed as a proof-of-concept demonstrating the utility of survival GWAS in developmental cohorts. We anticipate that the power of this approach will increase substantially when applied to larger datasets with longer follow-up periods and more diverse populations. Expanding analyses to cohorts with millions of individuals and broader ancestry representation will be critical for identifying robust and replicable genetic associations.

Several limitations should be considered. First, the sample size within each ancestry group remains modest, and the cohort is not enriched for substance use outcomes, resulting in limited event counts for some traits. Second, the relatively short follow-up period in ABCD may lead to incomplete capture of initiation events. Third, phenotypes are primarily based on self-report, which may introduce measurement error or misclassification. Finally, cross-ancestry meta-analysis may have reduced power in the presence of heterogeneity in genetic effects across populations. Future work should aim to extend follow-up duration, incorporate more refined and objective phenotyping, and apply trans-ethnic methods that explicitly model ancestry-specific genetic architecture. Replication in independent cohorts, such as the UK Biobank [32] or other longitudinal datasets, will also be important to validate these findings. Although access to such datasets is currently limited, we aim to address this in future studies.

## Conclusion

In summary, survival GWAS in the ABCD cohort identifies suggestive genetic signals associated with substance use initiation and demonstrates the feasibility of a strand-aware multi-ancestry meta-analysis framework. Our findings suggest partial shared genetic liability between alcohol and overall substance use initiation, alongside more trait-specific genetic architectures for nicotine and cannabis at the current sample size.

These results provide an important foundation for future studies integrating genetic, environmental, and developmental factors, as well as for downstream functional annotation and biological interpretation.

## Author Contributions

Mengman Wei conceived the study, designed the analytical framework, performed all data processing, statistical analyses, and computational modeling, and drafted the manuscript. All code implementation, data curation, and result interpretation were conducted by Mengman Wei.

Qian Peng provided supervision, general guidance, resource support, and funding acquisition.

## Preprint Notice

This manuscript is a preprint and has not yet undergone peer review. The content is shared to disseminate findings and establish precedence. Additional analyses and revisions may be incorporated in future versions.

## Data availability

### Code

The analysis code and scripts used in this study are freely available at the following GitHub repository: https://github.com/mw742/Survival-GWAS.

### Data

This study uses data from the Adolescent Brain Cognitive Development (ABCD) Study (https://abcdstudy.org), held in the NIMH Data Archive (NDA). The ABCD data release used was version 5.1. The study is supported by the National Institutes of Health (NIH) and additional federal partners under multiple award numbers, including U01DA041048 and U01DA050987. The full list of funders is available at https://abcdstudy.org/federal-partners.html.

